# Mathematical modeling of vaccination rollout and NPIs lifting on COVID-19 transmission with VOC: a case study in Toronto, Canada

**DOI:** 10.1101/2021.08.11.21261932

**Authors:** Elena Aruffo, Pei Yuan, Yi Tan, Evgenia Gatov, Iain Moyles, Jacques Bélair, James Watmough, Sarah Collier, Julien Arino, Huaiping Zhu

**Author notes:** Corresponding Author, 4700 Keele Street, Toronto, Ontario, Canada, M3J1P3. Co-first authors made the same contributions (Elena Aruffo, Pei Yuan, Yi Tan).

## Abstract

**Background:** Since December 2020, public health agencies have implemented a variety of vaccination strategies to curb the spread of SARS-CoV-2, along with pre-existing Nonpharmaceutical Interventions (NPIs). Initial strategy focused on vaccinating the elderly to prevent hospitalizations and deaths. With vaccines becoming available to the broader population, we aimed to determine the optimal strategy to enable the safe lifting of NPIs while avoiding virus resurgence.

**Methods:** We developed a compartmental deterministic SEIR model to simulate the lifting of NPIs under different vaccination rollout scenarios. Using case and vaccination data from Toronto, Canada between December 28, 2020 and May 19, 2021, we estimated transmission throughout past stages of NPI escalation/relaxation to compare the impact of lifting NPIs on different dates on cases, hospitalizations, and deaths, given varying degrees of vaccine coverages by 20-year age groups, accounting for waning immunity.

**Results:** We found that, once coverage among the elderly is high enough (80% with at least one dose), the main age groups to target are 20-39 and 40-59 years, whereby first-dose coverage of at least 70% by mid-June 2021 is needed to minimize the possibility of resurgence if NPIs are to be lifted in the summer. While a resurgence was observed for every scenario of NPI lifting, we also found that under an optimistic vaccination coverage (70% by mid-June, postponing reopening from August 2021 to September 2021can reduce case counts and severe outcomes by roughly 80% by December 31, 2021.

**Conclusions:** Our results suggest that focusing the vaccination strategy on the working-age population can curb the spread of SARS-CoV-2. However, even with high vaccination coverage in adults, lifting NPIs to pre-pandemic levels is not advisable since a resurgence is expected to occur, especially with earlier reopening.

## BACKGROUND

After months of implementation of nonpharmaceutical interventions (NPIs including school/business closures, physical distancing, and mask-wearing), to control the spread of SARS-CoV-2, in December 2020 many countries finally initiated vaccination campaigns^1,2,3^. The most recommended strategy was prioritizing the elderly and high-risk populations, followed by essential workers, and then the general public^4, 5, 6^. At the initial stage of vaccine distribution, strict NPIs were kept in place to avoid potential virus resurgence. Now, after almost six months of immunization, there is a need to establish an optimal vaccination strategy in order to safely lift NPIs while avoiding virus resurgence.

There have been numerous mathematical models aiming to identify the best vaccination strategy ^7, 8, 9, 10, 11,12, 13,14,15,16^. Early models focused on reducing the spread of the infection and identifying priority groups for receiving the first dose. For example, Matrajt et al^16^ developed a compartmental model with age structure to determine which age group should be vaccinated first. They showed that with low coverage, the elderly (60+ years) must be prioritized to reduce the number of deaths. Bubar et al^9^ showed that prioritizing younger ages (20-49 years) can reduce cumulative cases independently from rollout speeds and coverages. In Fall 2020, new, more transmissible, and virulent, variants of concern (VOC) were discovered^17,18,19,20^. In many areas of the world, VOC cases increased rapidly in the following months and became dominant over SARS-CoV-2 wildtype ^20, 21,22,23^. To that end, Giordano et al.^8^ investigated the impact of mass vaccination campaigns and NPI lifting while considering increased transmission due to VOCs. They found that NPI implementation is crucial even after the introduction of vaccination in the population. Moore et al.^11^ also introduced VOC in their age-structured model as well as vaccination and different levels of reopening. They similarly confirmed that relaxing NPIs too early will result in virus resurgence and noted the infeasibility of reaching herd immunity through vaccination. While vaccines have not yet been approved in young children, recent studies consider younger age groups when defining the best vaccine rollout for minimizing infections, hospitalizations, and deaths^9, 25, 26^. Meehan et al.^25^, for example, showed that prioritizing individuals aged 30-59 reduces the transmission, while prioritizing ages 65+ reduces deaths. They also found that when coverage is close to herd immunity, vaccinating middle-aged adults should be the prioritized, since young teenagers and children appear to have minimal impact. Shim et al.^26^ conducted an optimization analysis on vaccination strategy, taking into account age groups and vaccine efficacy, and found that for a vaccine with at least 70% efficacy, targeting ages 20-49 is best for reducing infections, and targeting ages 50+ is best for reducing mortality. Although existing studies provide important information for decision making, they do not capture the nuances around the impact on VOCs on vaccination efforts by age group in terms of both transmission and virulence, as well as vaccine effectiveness, in order to assess scenarios for the safe lifting of NPIs.

In this paper, we aimed to determine an optimal vaccination strategy to enable the safe lifting of NPIs while avoiding virus resurgence, using Toronto, Canada as a case study. We have extended the basic SIR compartmental model to reflect a variety of infectious and recovered states and incorporated age structure and vaccine status. We further included two strains of the virus, differentially affecting transmission, virulence, and vaccine effectiveness. We then assessed different reopening strategies given varying degrees of vaccine coverage by age group aiming to reduce infections, hospitalizations, and deaths.

## METHODS

### Data, model structure and assumptions

Our model is applicable to any geographical region where sufficiently detailed data are available. To study COVID-19 vaccination rollout and reopening strategies, we used data from Toronto, Canada between December 28, 2020 and May 19, 2021. To calibrate model parameters, we used data on cases, deaths, hospitalizations, and daily vaccine doses. We developed a compartmental model with age structure and vaccine status and extended the basic Susceptible-Infected-Recovered (SIR) framework to reflect additional disease states and two strains of the virus.

As of May 5, 2021, the Canadian government approved the use of the Pfizer-BioNTech COVID-19 vaccine in teenagers aged above 12 years ^27^. We divided the population into 6 age groups: 0-9, 10-19, 20-39, 40-59, 60-79, 80+ years (0-9 years are not considered for vaccination at this time but are important for transmission consideration). We extended the SIR disease states to further include latent, asymptomatic, and symptomatic infections, as well as population movement into hospitalization, recovery, or death states (i.e., SLAIHDR model, **Figure 1**). We incorporated two strains of the virus: wildtype and VOC (specifically, the B.1.1.7 variant, most commonly circulating at the time of model parametrization).

**Figure 1:**
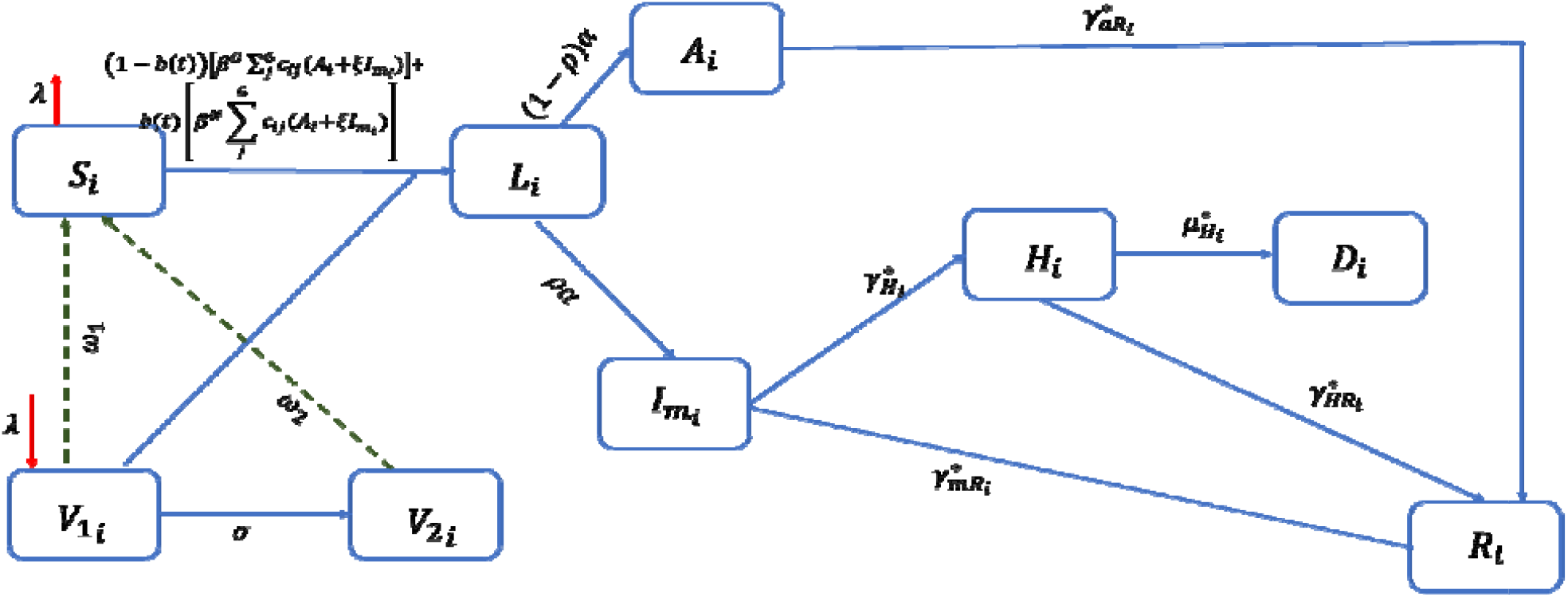
Flow diagram of COVID-19 transmission dynamics with two vaccination processes. Acronyms: i ∈{1-6}Age groups: 0-9 (unvaccinated), 10-19, 20-39, 40-59, 60-79, 80+; In Age group *i*: S_i_ (Susceptible), L_i_ (Latently infected), A_i_ (Asymptomatic infected), I_mi_ (Symptomatic mild infected), H_i_ (Hospitalized), D_i_ (Deceased), R_i_ (Recovered), V_1i_ (Vaccinated with first dose), V_2i_ (Vaccinated with second dose). To capture the different infection severities coming from VOC or wildtype variant, each disease-state progression is variant-dependent (*= wildtype or VOC). Red arrows: vaccination process. Dashed lines: waning process. Model assumptions:

- Only susceptible individuals, aged 10+ years, will receive the vaccine. Vaccine reduces susceptibility. Partially vaccinated people can become infected and infectious if the vaccine is not efficient
- Immunity follows two steps: partial (receiving 1 dose) and full (receiving 2 doses), with the second dose given after 112 days (in some predictive scenarios after 50 or 21 days)^33^. Immunity from one dose wanes in 120 days and from two doses after 365 days. Vaccination continues until 80% of the entire population receive at least one dose.
- Vaccine efficacy is age dependent (higher for teenagers and adults, 10% lower for elderly) and is the same against wildtype variant and VOC (all non-wildtype cases are assumed to be B.1.1.7 variant)
- VOC and wildtype are both included in the transmission process, assuming that the proportion of VOC cases increases over time following a sigmoidal function, with transmission from VOC 1.5x higher than wildtype
- Only individuals hospitalized might die from the infection

The infection dynamic is presented in **Figure 1**. The susceptible compartment (S), with age dependent susceptibility, can become infected with either the wildtype or VOC (indicated with O and N, respectively), with age dependent transmission rate *β c*_*ij*_, where c_ij_ is the number of contacts of individuals in age group *i* with individuals in age groups j and *β* is the probability of infection per contact. Multiple studies confirmed that VOC is roughly 40% to 90% more transmissible than the wildtype variant [29, 30, 31], hence we assumed that the probability of transmission of VOC (*β*^*N*^) is *ζ* times higher than the probability of transmission of the wildtype variant (i.e., *β*^*N*^ = *ζ β*^*o*^). Upon infection, individuals enter a latent stage (compartment E, exposed) where they are neither symptomatic nor infectious. We assume latently infected individuals become infectious at rate *α*: a fraction *ρ* develops mild symptoms (compartment I_m_) and the remainder remain asymptomatic for the duration of infection (compartment A). From A, individuals recover at rate *γ*_*aR*_. We further assume individuals with mild symptoms recover, at rate *γ*_*mR*_, or progress to clinical stage, at rate *γ*_*H*_, and reduce their contact rate (compartment H). In H, individuals can either recover, at rate *γ*_*HR*_, or become deceased (D) at rate *µ*_*H*_. To better describe the daily increase of cases coming from the VOC, we modelled the growth of cases from the novel variant using a sigmoidal function (**Figure SI1**). Each compartment is divided into 1 − *b*(*t*) proportion coming from the wildtype variant and b(t) proportion coming from the VOC. This allowed us to capture the differences between these variants in terms of probabilities of transmission and probabilities of severe outcomes. The model’s equations and parameters are shown in **SI Eq. SI1 and Table SI2**.

The population is further structured by vaccination status (none, partial and full), with no possibility of reinfection. The vaccine we chose to model has the characteristics of Pfizer/Moderna in that it is delivered in two doses^32^. Therefore, individuals move to *V*_1_ after receiving the first dose and *V*_2_ after receiving the second dose, where they are considered as fully immunized. We assumed that eventually all recipients of the first dose will be vaccinated with the second one and this occurs at a rate *σ*. Vaccine efficacy, *ε*, of both doses is included and assumed the same against VOC and wildtype variants. We assumed that vaccine efficacy reduces the probability of infection (*β c*_*ij*_,) by 1 − *ϵ*. Immunity induced by vaccination is assumed to wane after one and two doses at different rates. In the model, we simulated a minimum vaccine coverage that each age group needs to reach by June 14, 2021. Thereafter, the vaccination process continues until 80% of the population is vaccinated.

### Reopening scenarios analysis

With increasing vaccination rollout, public health has considered easing some NPI restrictions^28^. Therefore, we predicted cumulative cases and deaths, and daily hospitalizations until December 31, 2021, comparing different degrees of reopening, at different dates, given a variety of vaccine coverages for each age group. Using historical case data and information on previous policy periods of escalating/de-escalating NPIs in Toronto, we have identified four distinct stages of transmission with varying degrees to which indoor/outdoor gatherings were permitted, retail at full, limited, or curbside-only capacity, and whether indoor/outdoor dining and other sectors, including cinemas and gyms, were open. Compared to the most recent level of restriction up to May 19, 2021, during which retail was curbside only, with no indoor/outdoor dining, stay-at-home in effect, and personal protective (PP) behaviours such as physical distancing and mask-wearing enforced, we included possible permutations for reopening on June 15, August 15, or September 15 to different degrees (1) No reopening (i.e., remain at baseline); (2) Partial reopening, whereby contacts are increased by 50% compared to baseline, reflecting a small increase in gatherings and retail; (3) Total reopening with contacts increased by 70% and the probability of transmission increasing by 35%, compared to baseline, reflecting most sectors being open with a more relaxed use of PP; and (4) Pre-pandemic contact rates with no limitations on gatherings and with all sectors being open, while still maintaining PP. Pre-pandemic contacts within and between age groups were calculated using contact matrices from Canada, as described elsewhere^34^ (for detailed calculations and matrices, see **SI and Table SI3**). For each permutation, we calculated the reproduction number *R*_*c*_ using the Next Generation Matrix method^35, 36^, and identified the vaccination coverage, by age, that would in *R*_*c*_ below 1.

For each reopening scenario, we examined the impact of vaccination by age group. We used vaccination data up to May 19 to estimate the vaccination rate required to reach specific coverages by June 14, 2021 (a plateau, or a 10%, 20% or 30% increase from current coverage for each age group), all the model permutations are given in **SI Figure SI3**. We then used the average daily doses from that day moving forward, until 80% of the entire vaccinable population has received at least one dose. Since the most recent vaccine coverage among ages 60+ was above 70%, we primarily focus on varying coverages in those under 60 years of age, assuming that by mid-June, ages 60-79 and 80+ might reach 80%-90% coverage with the first dose, ages 40-50 might reach 70%-90%, ages 20-39 might reach 60%-80%, and ages 10-19 might reach 20%-40%. Given guidelines on extended timeframes with limited vaccine supply [33], we assumed that the second dose is given 112 days later, also compared scenarios with a shorter interval between doses (50 or 21 days). The first and second dose ere assumed to be 90% and 80% effective, respectively, with 10% reduction in effectiveness among ages 80+ (see **SI Table SI2**).

### Sensitivity Analysis

To explore the impact that vaccination coverages and time between doses have on the model outcomes, we investigated a sensitivity analysis of these parameters using the Latin Hypercube Sampling-Partial Rank Correlation Coefficient (LHS-PRCC) method. We generated 1000 samples using the LHS method with uniform distribution and investigated correlation between the samples and model outputs, such as cumulative cases and deaths. The scenario used is total reopening in June. Parameters with a PRCC whose absolute value is larger than 0.5 are considered significant. Ranges of sampled parameters are shown in **SI Table SI2**.

## RESULTS

### Reproduction number

We investigated the reproduction number considering that the coverage of the age groups 10-19, 60-79 and 80+ years is 20%, 80%, 90%, respectively, and varying the coverages for the remaining groups from 50% to 90%. The model employed for this study is data-driven and control strategies dependent and we observe that the system does not have a disease-free state. Moreover, the susceptible compartment is reduced daily by a time-depended vaccination rate, reflecting the daily doses given to each age group. We investigated the reproduction number by looking at the total coverage that age groups might achieve, reducing the susceptible class by this proportion and adding it to the vaccinated compartment.

We observe that, if current contacts are increased by 50% and PPEs are in place, the reproduction number remains below 1 if both age groups 20-39 and 40-59 years reach a minimum coverage of 60% (see **SI Figure SIA**). On the other hand, if contacts are increased by 70% and the probability of transmission is increased by 35% (see **SI Figure SI4B**), the reproduction number will never be below 1, unless more than 90% of adults are vaccinated. **Figure SI4C** shows that with an NPIs pre-pandemic reopening, the infection will continue spreading with the highest reproduction number.

### Prediction of best reopening strategies and vaccination coverages

The tables in the following sections represent the percentage change of the shown scenario with respect to the baseline case, defined as the minimum coverages of each sub-populations and no reopening scenario (see **SI Figure SI3**).

### Identification of age group that minimizes cases, deaths and hospitalizations

Table 1 shows the percentage change of cumulative cases by the end of December 2021, with respect to the base line in **SI Figure SI3**, when partial reopening occurs on September 15. When efforts are mainly put into vaccinating the 10-19 years age group, the change in cases does not appear to be significant. For example, with a 60% coverage of the 20-39 age group, if the youngest group is vaccinated from 20% to 40%, cases are reduced by roughly 7%. However, increasing the coverage of the 20-39 age group by 20% will result in reducing cases by roughly 70%, respectively. On the other hand, if the youngest age group is at minimum coverage (i.e., 20%), the cumulative cases remarkably decrease as the coverage of age groups 20-39 years and 40-59 years increase. We observe that if 20-39- and 40-59-years groups are vaccinated above 80%, respectively, the increase from the baseline varies between 2.47% and 5.98%. If the 20-39 age group reaches 80% coverage, then increasing the coverage of the 40-59 age group from 60% to 70% or 80% reduces cases by 49% and 79%, respectively. If the coverage of age group 20-39 years is reduced to 60%, the increase can reach 43.6%.

**Table 1:** Percentage change of cumulative cases with respect to the base line NPIs no reopening in SI Figure SI3 with total reopening in September, when age groups 60-79 and 80+ reached coverages 80%, 90% by June 14. Cases are reported comparing different coverages for age group 10-19 years, assuming 40-59 years fixed at 70% coverage (top table) and comparing different coverages for age group 40-59 years, assuming 10-19 years fixed at 20% coverage (bottom table). The second dose is given at a rate of 1/112 days^-1^

| Projected percentage change of <u>cumulative cases</u> with respect to baseline NPIs no reopening in SI Figure SI3 after reopening in September with NPIs total reopening |  |  |  | Projected percentage change of <u>cumulative deaths</u> with respect to baseline NPIs no reopening in SI Figure SI3 after reopening in September with NPIs total reopening |  |  |  |
| --- | --- | --- | --- | --- | --- | --- | --- |
|  |  | 20-39 years coverage by June 14, 2021 |  |  |  | 20-39 years coverage by June 14, 2021 |  |
|  |  | 60% | 80% |  |  | 60% | 80% |
| 10-19 years coverage by June 14, 2021 | 20% | 43.6 | 11.85 | 10-19 years coverage by June 14, 2021 | 20% | 33.4 | 10.67 |
|  | 30% | 42.2 | 10.81 |  | 30% | 32.44 | 9.88 |
|  | 40% | 40.5 | 11.73 |  | 40% | 31.22 | 10.63 |
|  |  | 20-39 years coverage by June 14, 2021 |  |  |  | 20-39 years coverage by June 14, 2021 |  |
|  |  | 60% | 80% |  |  | 60% | 80% |
| 40-59 years coverage by June 14, 2021 | 70% | 43.6 | 11.85 | 40-59 years coverage by June 14, 2021 | 70% | 33.4 | 10.67 |
|  | 80% | 26.11 | 5.98 |  | 80% | 20.8 | 6.3 |
|  | 90% | 14.46 | 2.47 |  | 90% | 12.36 | 3.69 |

Our results also show that, if age group 20-39 years is highly vaccinated, immunizing more than 30% of teenagers is not beneficial, since we observe an increase in cases (for example 10.81% with 30% coverage to 11.73% with a 40% coverage). This result is due to the fact that after June 14 the vaccination process continues until the total vaccinated population reaches 80%. If we increase the vaccination rate of the 10-19 age group, the total coverage is reached earlier leaving some age groups still susceptible. In particular, the age group 40-59 years will not reach enough coverage to prevent the increase of cases.

The results for cumulative deaths are similar to those for cumulative cases. Since the elderly population is already highly vaccinated, it is important to focus on the immunization of the age groups 20-39 and 40-59 years reach at least 80% coverage to reduce deaths after reopening.

**Figure 2** shows hospitalizations under the scenario of NPIs total reopening in September, if 60%-80% of the 20-39 age group is vaccinated and the 10-19 age group coverage is 20%-40% (**A**) or the 40-59 age group is 70%-90% (**B**). Like Table 1, we observe that there is no significant reduction in the number of hospitalizations if the coverage of the youngest age group is increasing from 20% to 40% (**Figure 2A**). However, if at least 80% of young adults are vaccinated, then hospitalizations drop by roughly 87% (blue line and red line in **Figure 2A**). On the other hand, analyzing different coverages for the 40-59 age group, we observe that if at least 80% of adults aged between 20 and 39 years are vaccinated and 90% of the 40-59 years population, the increase of hospitalizations is much lower than the scenario with minimum coverage (dashed purple line vs blue line, **Figure 2B**). In both analyses, we observe an increasing trend of hospitalizations after the reopening, suggesting that an NPIs total reopening strategy is not beneficial.

**Figure 2:**
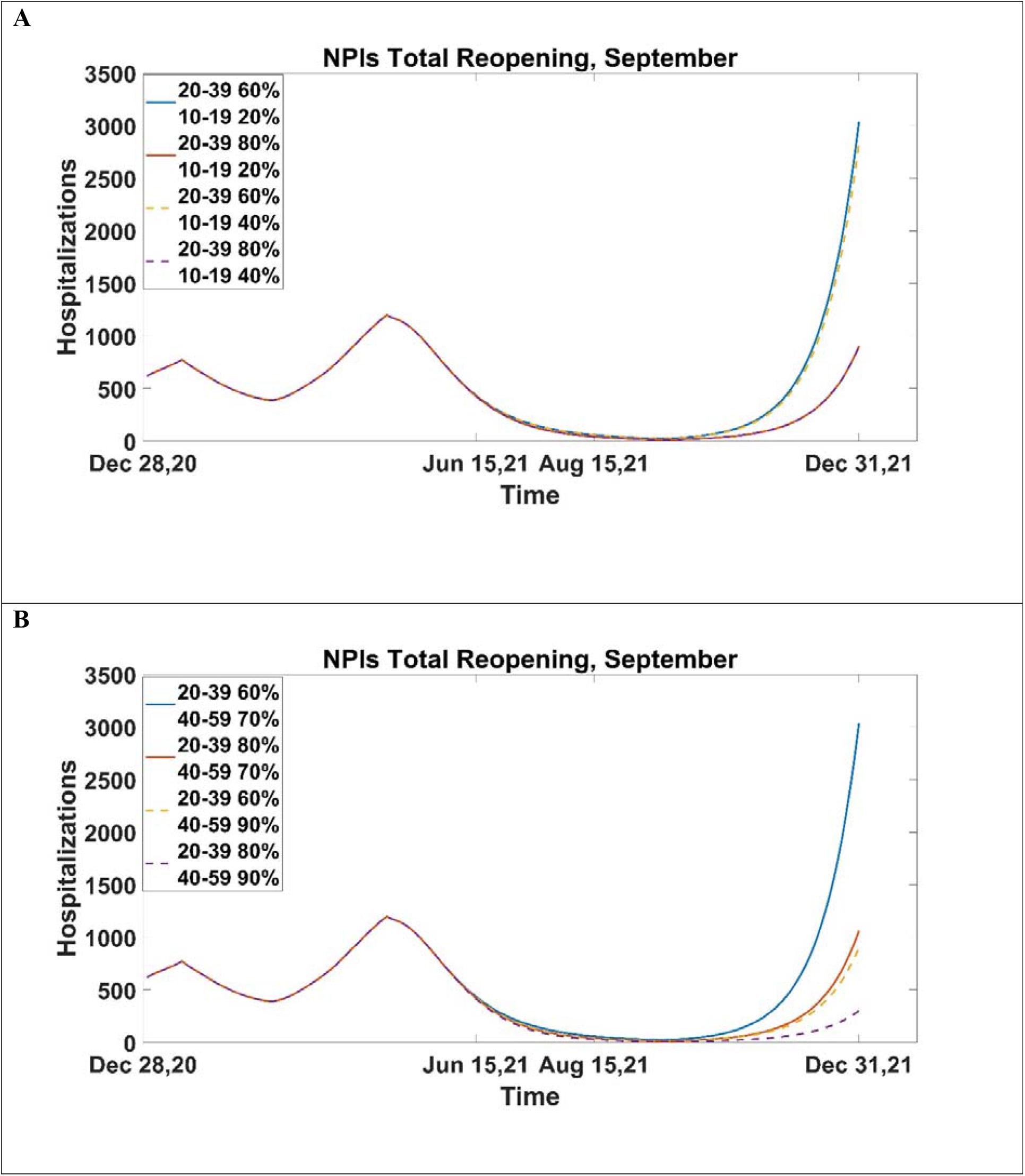
Hospitalizations with partial reopening in September (A) if 10-19 is vaccinated 20%-40%, 20-39 60%, 80% and 40-59, 60-79 and 80+ reached coverages 70%, 80%, 90%; (B) if 40-59 is vaccinated 70%-00%, 20-39 60%, 80% and 10-19, 60-79 and 80+ reached coverages 20%, 80%, 90%. The second dose is given at a rate of 1/112 days^-1^.

### Identification of the best combination of vaccination coverages and NPIs lift levels

From Table 2, we immediately observe that if NPIs partial and total reopening occur in August, cases increase up to 235% from the baseline, reaching a 29% increase with the highest vaccination coverage in the age groups 20-39 and 40-59 years. On the other hand, we observe that with NPIs pre-pandemic reopening, reopening earlier is slightly more beneficial than reopening later with the lowest coverage among adults. This is due to the assumption of a fast-waning immunity rate for partially immunized individuals and suggests that in this scenario, if reopening occurs later, low coverages and more individuals becoming susceptible within the period of pre-reopening, the infection spreads once NPI’s are lifted completely.

**Table 2:** Percentage change of cumulative cases with respect to the baseline with respect to the base line NPIs no reopening in SI Figure SI3 with partial, total and pre-pandemic reopening in August and September, when age groups 10-19, 60-79 and 80+ reached coverages 20%, 80%, 90%. The second dose is given at a rate of 1/112 days^-1^.

| Projected percentage change of <u>cumulative cases</u> with respect to the base line NPIs no reopening in SI Figure SI3 |  |  |  |  |  |  |  |
| --- | --- | --- | --- | --- | --- | --- | --- |
|  |  | In reopen in AUGUST |  |  |  |  |  |
|  |  | 20-39 years<br>coverage by June 14, 2021 |  |  |  |  |  |
|  |  | 60% |  |  | 80% |  |  |
| NPI's Level of reopening |  | Partial | Total | Pre-pandemic | Partial | Total | Pre-pandemic |
| 40-59 years<br>coverage by June 14,<br>2021 | 70% | 3.56 | 235 | 932 | -0.004 | 81.5 | 894 |
|  | 80% | 1.72 | 162 | 916 | -0.86 | 49.8 | 877 |
|  | 90% | 0.36 | 103 | 898 | -1.57 | 29 | 860 |
|  |  | In reopen in SEPTEMBER |  |  |  |  |  |
|  |  | 20-39 years<br>coverage by June 14, 2021 |  |  |  |  |  |
|  |  | 60% |  |  | 80% |  |  |
| NPI's Level of reopening |  | Partial | Total | Pre-pandemic | Partial | Total | Total |
| 40-59 years<br>coverage by June 14,<br>2021 | 70% | -8.18 | 43.6 | 938 | -8.71 | 11.85 | 874 |
|  | 80% | -8.45 | 26.11 | 915 | -8.9 | 5.98 | 835 |
|  | 90% | -8.69 | 14.46 | 885 | -9.1 | 2.47 | 783 |

We also observe that if the reopening is NPIs partial, then the reduction in cases given by different vaccination strategies is not significant (8.18% to 9.1%) if the reopening occurs in September. This suggests that a low level of reopening helps in controlling the spread of the virus. On the other hand, with the same NPI lift level occurring a month earlier, cases increase with respect to the baseline if there is low vaccine coverage in adults. However, if the coverage of 20-39 and 40-59 years achieve 80%, then we see a reduction of cases once more with the highest coverage providing a reduction of 144% of cases. If the NPI lift levels are higher (total or pre-pandemic) in September, then we observe a reduction in cases as the vaccination coverage increases in the adult groups. However, the impact of vaccination coverage appears to be more significant with NPIs total reopening. For example, a 20% increase in coverage of 20-39 years is roughly 6%-11% with NPIs pre-pandemic reopening, and 73%-82% with NPIs total reopening. Similar trend is observable if reopening occurs in August.

The projections of cumulative deaths show similar results of cumulative cases (see **SI Table SI4**). However, we observe that, if NPIs are lifted with NPIs partial in August and adults vaccinated above 70%-80%, the percentage change with respect to the baseline is negative for cumulative cases, while it is positive for the projected cumulative deaths. Also, with pre-pandemic reopening, the percentage change of deaths is higher than the cases, suggesting that for a pre-pandemic reopening, vaccinating adults is not enough to reduce deaths.

Comparing **Figure 2B** and **Figure SI5**, we observe that if the community is reopened with partial reopening in August, rather than in September, the hospitalizations increase by five times. Even with the highest coverages among adults, with reopening in August, the hospitalizations reach 2185 by the end of 2021 against the 360 projected, by the same date, if the reopening occurs in September.

### Identification of the best combination of vaccination coverages and NPIs lift date, with lowest efficacy

**Table 3** presents the percentage change of cumulative cases and deaths under different NPI lift levels and dates, with the vaccine efficacy against the virus reduced by 10%. We observe that a lower efficacy leads to a large increase of cases and deaths (up to 200% of increase). The highest reduction in cases is given with total NPIs reopening and when the coverage of age groups 20-39 and 40-59 years exceeds 60% and 70% respectively. A coverage of 80% and 90% in these two groups provides the lowest increase from the baseline. With a stricter reopening, the efforts in reaching these coverages are not evidently beneficial; in fact, cases are reduced by roughly 2%.

**Table 3:**
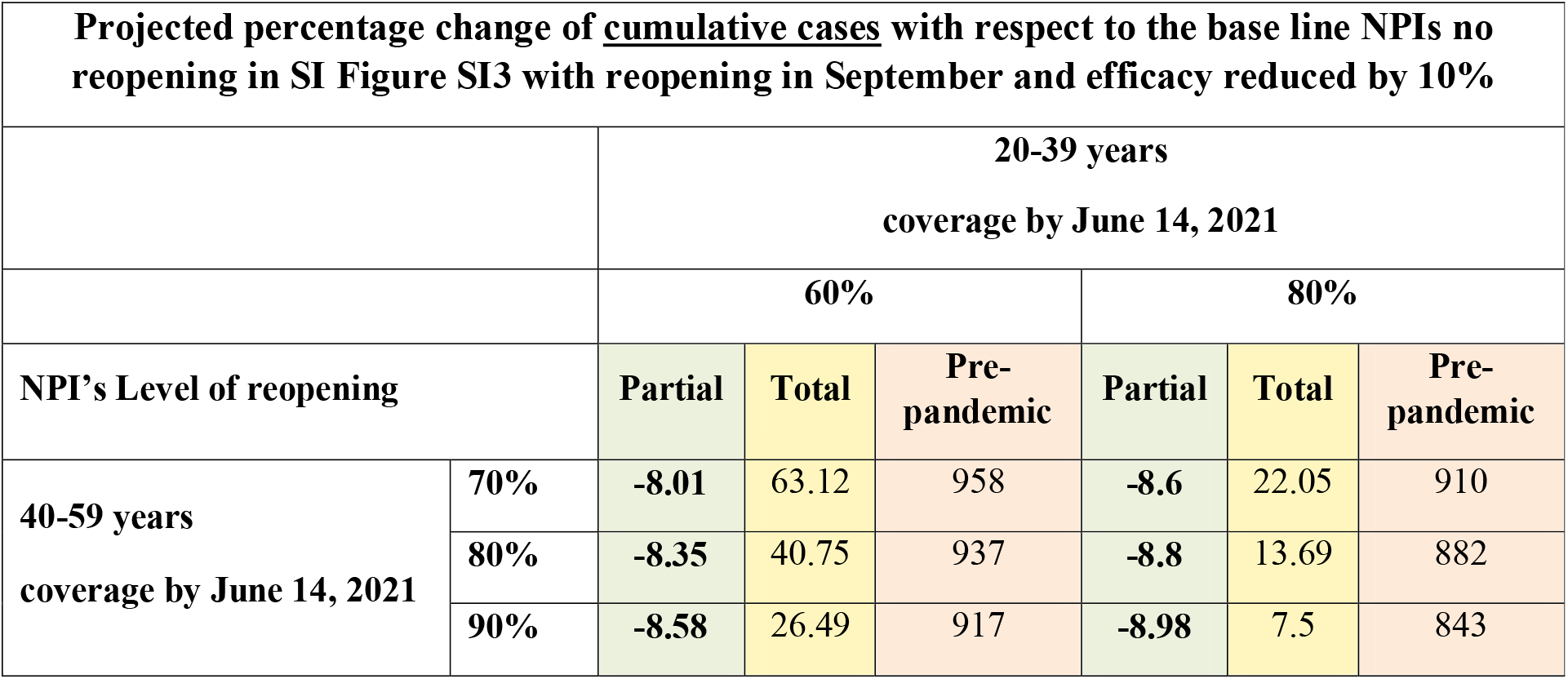
Percentage change of cumulative cases with respect to the baseline with respect to the base line NPIs no reopening in SI Figure SI3, reducing efficacy by 10%, with partial, total and pre-pandemic reopening when age groups 10-19, 60-79 and 80+ reached coverages 20%, 80%, 90%.

**Figure 3** shows hospitalizations until the end of 2021. A reduction of 10% in efficacy will result in an increase of hospitalizations from 3038 to 5515 with low coverage of vaccination and from 302 to 692 with highest coverage at the end of December 2021.

**Figure 3:**
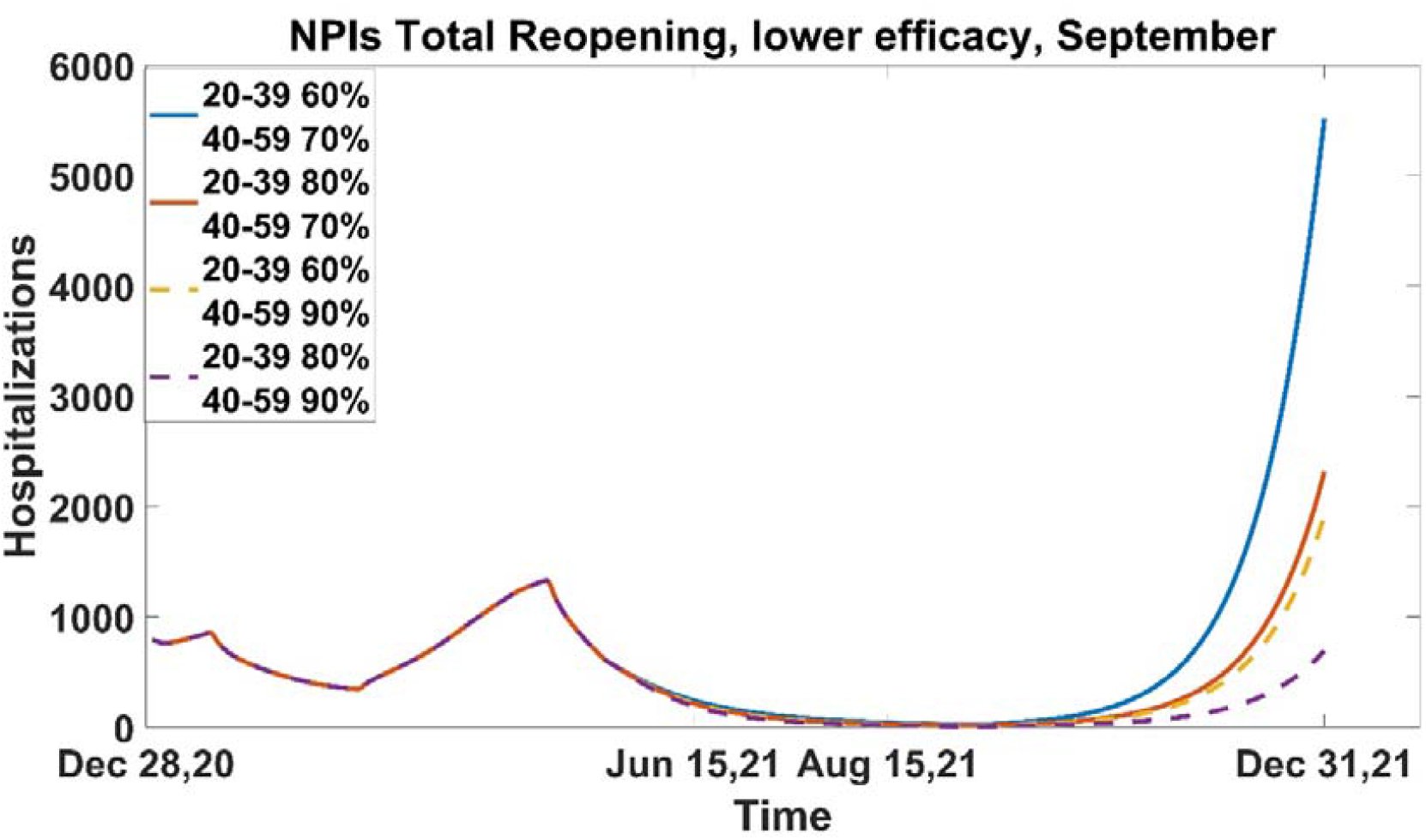
Hospitalizations if 40-59 is vaccinated 70%-90%, 20-39 60%, 80% and 10-19, 60-79 and 80+ reached coverages 20%, 80%, 90% with total NPIs reopening in September with efficacy decreased by 10%.

The percentage change of cumulative deaths is reported in Table A4. We observe that even for cumulative deaths, the highest reduction is possible with any coverage as long as the level of reopening is One. On the other hand, if the level increases, then it is necessary to increase the vaccination coverage among adults above 80%.

### Effect of reducing time between first and second dose

Until the end of May 2021, in Ontario the second dose of vaccine was given after 16 weeks from the first one. However, after June, this timeframe was shortened to 12 weeks^37^. According to the recommendation provided by the pharmaceutical manufacturing companies the second dose should be given after 21 days from the first one^32^. We compared how shortening the time needed to reach full immunization impacts the spread of the infection (see **Table 4**).

**Table 4:** Percentage change of cumulative cases with respect to the base line NPIs no reopening in SI Figure SI3 with partial, total and pre-pandemic reopening in September and second dose given after 21 or 50 days. Age groups 10-19, 60-79 and 80+ are assumed to reach coverages 20%, 80%, 90% by mid June. Par.= partial; Tot.= total; Pre-pan.= pre-pandemic.

| Projected percentage change of <u>cumulative cases</u> with respect to the base line NPIs no reopening in SI Figure SI3 with reopening in September |  |  |  |  |  |  |  |  |  |  |  |  |  |
| --- | --- | --- | --- | --- | --- | --- | --- | --- | --- | --- | --- | --- | --- |
|  |  | 21 days |  |  |  |  |  | 50 days |  |  |  |  |  |
|  |  | Between dose 1 and dose 2 |  |  |  |  |  | Between dose 1 and dose 2 |  |  |  |  |  |
|  |  | 20-39 years<br>coverage by June 14, 2021 |  |  |  |  |  | 20-39 years<br>coverage by June 14, 2021 |  |  |  |  |  |
|  |  | 60% |  |  | 80% |  |  | 60% |  |  | 80% |  |  |
| NPI's Level of reopening |  | Par. | Tot. | Pre-pan. | Par. | Tot. | Pre-pan. | Par. | Tot. | Pre-pan. | Par. | Tot. | Pre-pan. |
| 40-59 years<br>coverage by June 14, 2021 | 70% | -8.7 | 4.56 | 753 | -9.1 | -1.24 | 488 | -8.4 | 15.5 | 848 | -8.9 | 2.05 | 719 |
|  | 80% | -8.9 | 1.03 | 662 | -9.3 | -2.19 | 320 | -8.6 | 7.96 | 805 | -9.1 | -0.13 | 621 |
|  | 90% | -9.1 | -0.92 | 535 | -9.5 | -2.77 | 173 | -8.86 | 3.22 | 745 | -9.2 | -1.38 | 491 |

Comparing **Table 4** to **Table 2** (bottom), we observe that a faster rollout of second dose is always beneficial. With pre-pandemic NPIs reopening and minimal coverage of age groups 40-59 years and 60% or 80% coverage of 20-39 years age group, we see that reducing the time between doses by almost half, cases will reduce by 9% and 17%, respectively. Moreover, if the second dose is given according to 21 days as suggested by the manufacturer, the reduction can further reach 19% and 44% respectively. We also note that for a total NPIs reopening, if fully immunization is reached after 112 days, cases increase even with the highest coverages among adults. On the other hand, if the vaccination coverage of 20-39 and 40-59 years exceeds 80%, then a reduction in cases varies from 102% and 155%, if a second dose is given after 50 days, and 136% and 212%, if full immunization is reached after 21 days. Also, we observe that with lowest coverage of adults, shortening the time between the two doses is still beneficial. For example, with 60% coverage and reducing the time from 50 to 21 days, cases are dropped by 70%.

Minimizing the time between doses is also advantageous to reduce the number of deaths and hospitalizations (see **SI Table SI6 and Figure SI6**). With the lowest coverage of vaccination among adults, reducing the time from 112 days to 21 days, deaths are reduced 17% to 68%. With a total NPIs reopening and minimum vaccine coverages, hospitalizations are decreased by 55.6% and 85% if the second dose is given after 50 or 21 days respectively instead of 112 days. The reduction will become 76.5% and 95.4% respectively under the highest vaccine coverages.

### Sensitivity Analysis

Sensitivity analysis conducted on the daily doses and rate at which the second dose is given shows that the parameters more significant on the cumulative cases and deaths, and hospitalizations 50 days after reopening are the vaccination rates of age groups 20-39 and 40-59 years and the time between doses (see **Table SI7**). In particular, the PRCC values show negative correlation between these parameters and the model outcomes. This result suggests that not only adults need to be targeted to reduce cases, deaths, and hospitalizations, but also reducing the time between doses is beneficial.

## DISCUSSION

We developed an age-structured compartmental model which captures the infection dynamics of COVID-19. The SLAIHDR model considers vaccination and waning processes and an infectious compartment that captures both symptomatic and asymptomatic cases. The number of hospitalized and deceased cases are also included. The population is divided into six groups and assumes that children aged 0 to 9 years are not immunized against the SARS-CoV-2 virus. Given the emergence of new variants, the growth of cases deriving from variants of concern (VOC) was captured using a time-dependent sigmoidal function. This needed to be included in the model to better predict the course of the infection and effectiveness of treatment. This approach can identify severity differences between strains for outcomes such as death and hospitalization rates.

Our analysis shows that prioritizing the age group 10-19 in the vaccination rollout will not provide a large impact in reducing cases, unless the adult groups are vaccinated above 80% by mid June 2021. In fact, reaching this coverage in the 20-39 and 40-59 age groups will maximize the reduction of cases, deaths and hospitalizations. Sensitivity analysis confirms this result. Our results also confirm, as expected, that a late partial and total NPIs reopening will reduce the infection outcomes by roughly 80%; we still observe that to minimize the infection spread, the proportion of adults that needs to be vaccinated by mid June is 80%. However, even if delayed, a complete reopening, with pre-pandemic contacts, will result in a visible spread of infection, also with the highest vaccine coverage.

As of June 14, 2021, the coverage of adults is 76.12% and 72.9% for the age groups 20-39 and 40-59 years, respectively. Hence, our results suggest that if vaccination rollout is not accelerated, a reopening might result in a higher resurgence of the infection.

With new variants circulating, the vaccine’s efficacy against them plays an important role in the vaccine rollout. Our analysis on the vaccine distribution and reopening strategies shows that with a lower efficacy against the virus, deaths increase by roughly 50% with NPI Level Two reopening in September compared to the high efficacy scenario.

The time at which NPIs are lifted has a great impact on the control of the infection. Our results show that in general, reopening in September, rather than August, is more beneficial. In fact, with partial and total NPIs reopening, cases increase by 100%-400% if the reopening occurs one month earlier. Reopening in August is beneficial only with partial NPIs reopening and the adult population vaccinated above 70%.

Since the second dose provides a higher efficacy, compared to the first one, a faster distribution of vaccine to reach full immunization can control the spread more quickly. In fact, our results on the second doses administered after 21 or 50 days, shows that there is a higher reduction of cases if full immunity is provided after 3 weeks from the first shot. This result is expected from the formulation of our model, since a shorter period between doses will increase the number of individuals who are fully immunized faster.

Our study has some limitations. Firstly, we assumed that all the VOC cases are coming from B1.1.7 and the efficacy against the virus is the same for wildtype variants and VOC. However, as new variants emerge, with a much lower vaccine efficacy, it will be important in future work to consider multiple strains to better capture the role of efficacy and vaccine rollout. Secondly, we assume that recovered individuals from any variant are not susceptible to other variants, but with stronger variants emerging, infection-acquired immunity might protect individuals only partially. Thirdly, all individuals vaccinated with the first dose will eventually receive the second dose, but awareness can decrease, and a fraction of people might decide not to receive the second dose. Lastly, we assumed that vaccination is effective since the day it is receives, even if recent studies show that full protection is reached roughly after two weeks from the inoculation.

## CONCLUSIONS

In conclusion, our model reflects the course of COVID-19 infection in Toronto considering infection from the VOC and original wildtype strain. We were able to capture, through data, the different infection outcomes such as transmission, hospitalizations, and deaths, generated by different variants of the virus. Our results show that it is imperative to direct our efforts towards individuals aged between 20 and 59 years, showing similarities with previous works^23, 6, 22^. In fact, these are the age groups with higher contacts, social activity, and population size. Moreover, we showed that if the reopening strategy includes a complete return to pre-pandemic contact, the infection will immediately increase, even with the highest coverages reached by mid June. Also, we showed that the introduction of new variants, the vaccine efficacy against them, and the reduced time to obtain full immunity play an important role in the vaccination rollout. A reduction in efficacy will lead to a higher spread of the infection, and if, additionally, the second dose is delayed too much, the risk of having a re-emergence of the infection is possible.

## Data Availability

All data used is available in the manuscript.

https://github.com/EAruffo/Covid_vaccine_NPIs_paper

## ARTICLE INFORMATION

## Acknowledgements

We would like to acknowledge Toronto Public Health for the use of case and vaccination data in this study.

## Authors’ contributions

Research design: H.Z., E.A., P.Y., Y.T., E.G., S.C; Literature search: E.A., P.Y., Y.T.; Data collection: E.A., P.Y., Y.T; Modeling: H.Z. and all; Model analysis: E.A., P.Y., Y.T.; Simulations: E.A., P.Y., Y.T; Draft preparation: E.A., P.Y., Y.T, H.Z.; Writing reviewing-editing: H.Z., E.A., P.Y, Y.T., E.G., I.M., J.B., J.W., S.C., J.A., Supervision: H.Z.

## Data availability

The datasets generated and/or analysed during the current study are available in the Covid_vaccine_NPIs_paper repository, https://github.com/EAruffo/Covid_vaccine_NPIs_paper. Parameters used to generate analyses are provided in Supplemetary Information.

