## Supplementary Information for "Mathematical modeling of vaccination rollout and NPIs lifting on COVID-19 transmission with VOC: a case study in Toronto, Canada"

### ***Model***

The system of ODEs describing the dynamic is given by:

$$
\begin{aligned}
S_i' &= -\lambda_{s_i} - \phi_i S_i \left\{ \left(1 - b(t)\right) \left[ \beta^O \sum_j^5 c_{ij}(A_i + \xi I_{m_i}) \right] \right. \\
&\qquad \left. - \; b(t) \left[ \beta^N \sum_j^5 c_{ij}(A_i + \xi I_{m_i}) \right] \right\} + \omega_1 V_{1i} + \omega_2 \mathrm{V_{2i}} \\
E_i &= \phi_i S_i \left\{ \left(1 - b(t)\right) \left[ \beta^O \sum_j^5 c_{ij}(A_i + \xi I_{m_i}) \right] - \; b(t) \left[ \beta^N \sum_j^5 c_{ij}(A_i + \xi I_{m_i}) \right] \right\} \\
&\qquad + \phi_i V_{1i}\left(1 - \epsilon_{1_i}\right) \left\{ \left(1 - b(t)\right) \left[ \beta^O \sum_j^5 c_{ij}(A_i + \xi I_{m_i}) \right] \right. \\
&\qquad \left. - \; b(t) \left[ \beta^N \sum_j^5 c_{ij}(A_i + \xi I_{m_i}) \right] \right\} - \left(1 - b(t)\right)\alpha^o E_i - b(t)\alpha^N E_i \\
A_i' &= (1 - \rho)\left(1 - b(t)\right)\alpha^o E_i + (1 - \rho) b(t) \alpha^N E_i - \; \left(1 - b(t)\right)\gamma_{aR_i}^O \mathrm{A_i} \\
&\qquad - b(t)\gamma_{aR_i}^N \mathrm{A_i} \\
I'_{m_i} &= \rho\left(1 - b(t)\right)\alpha^o E_i + \rho b(t) \alpha^N E_i - \left(1 - b(t)\right)\gamma_{H_i}^o \mathrm{I_{m_i}} - b(t)\gamma_{H_i}^N \mathrm{I_{m_i}} \\
&\qquad - \left(1 - b(t)\right)\gamma_{mR_i}^O \mathrm{I_{m_i}} - b(t)\gamma_{mR_i}^N \mathrm{I_{m_i}} \\
H_i' &= \left(1 - b(t)\right)\gamma_{H_i}^o \mathrm{I_{m_i}} + b(t)\gamma_{H_i}^N \mathrm{I_{m_i}} - \left(1 - b(t)\right)\mu_{H\,i}^o H_i - \; b(t)\mu_{H\,i}^N H_i \\
&\qquad - \left(1 - b(t)\right)\gamma_{HR_i}^O H_i - b(t)\gamma_{HR_i}^N H_i \\
D_i'^o &= \left(1 - b(t)\right)\mu_{H\,i}^o H_i + \; b(t)\mu_{H\,i}^N H_i \\
R_i' &= \; (1 - b(t))\gamma_{aR_i}^O \mathrm{A_i} + b(t)\gamma_{aR_i}^N \mathrm{A_i} + \left(1 - b(t)\right)\gamma_{mR_i}^O \mathrm{I_{m_i}} + b(t)\gamma_{mR_i}^N \mathrm{I_{m_i}} \\
&\qquad + \left(1 - b(t)\right)\gamma_{HR_i}^O H_i + b(t)\gamma_{HR_i}^N H_i \\
V_{1i} &= \lambda_{s_i} - \phi_i V_{1i}\left(1 - \epsilon_{1_i}\right) \left\{ \left(1 - b(t)\right) \left[ \beta^O \sum_j^5 c_{ij}(A_i + \xi I_{m_i}) \right] \right. \\
&\qquad \left. - \; b(t) \left[ \beta^N \sum_j^5 c_{ij}(A_i + \xi I_{m_i}) \right] \right\} - \sigma \epsilon_{2_i} V_{1i} - \omega_1 \mathrm{V_{1i}} \\
V_{2i} &= \sigma \epsilon_{2_i} V_{1i} - \omega_2 \mathrm{V_{2i}}
\end{aligned}
$$

Eq.SI1

For $i \in \{1,2,3,4,5,6\}$, where $\beta^N = \zeta \beta^O$.

The list of variables and assumptions is given in Table SI1.

**Table SI1:** Table of the model's variables and assumptions

| **Variable** | **Definition** |
|---|---|
| $S_i$ | Susceptible individuals in age group i |
| $L_i$ | Latently infected individuals in age group i |
| $A_i$ | Asymptomatic individuals in age group i |
| $I_{m_i}$ | Symptomatic (mild) individuals in age group i |
| $H_i$ | Hospitalized individuals in age group i |
| $D_i$ | Deceased individuals in age group i |
| $R_i$ | Recovered individuals in age group i |
| $V_{1_i}$ | Vaccinated individuals in age group i (first dose) |
| $V_{2_i}$ | Vaccinated individuals in age group i (second dose) |
| $i \in \{1,2,3,4,5,6\}$ | Age groups: 0-9,10- 19, 20-39, 40-59, 60-79, 80+ years respectively |

| **Assumptions** |
|---|
| 1. Only susceptible individuals, aged 10 years and older, will receive the vaccine |
| 2. Immunity follows two steps: partial (receiving one dose) and full (receiving two doses) |
| 3. The vaccine efficacy is age dependent (higher for teenagers and adults, lower for elderly) |
| 4. The vaccine efficacy is the same against wildtype variant and VOC |
| 5. The second dose is given after 112 days (in some predictive scenarios after 50 or 21 days), following the suggestion announced by the Government of Ontario in March 2021 [33] |
| 6. Immunity wanes from one dose of vaccine after 120 days and from two doses after 365 days |
| 7. We assume that the coverages in Table 2 are reached by June 14, 2021, and continue the vaccination process until 80% of the total population is vaccinated |
| 8. We assume that all non-wild type cases belong to B.1.1.7 |
| 9. VOC and wildtype are both included in the transmission process, assuming that proportion of cases from VOC increases by time, following a sigmoidal function |
| 10. The transmission from VOC is assumed to be 1.5 higher than the original variant |
| 11. Vaccine reduces susceptibility. Partially vaccinated people can become infected and infectious if the vaccine is not efficient |
| 12. Only individuals hospitalized might die from the infection |

**Table SI2**: Table of model parameters

| Parameter | Definition | Value | Ref. |
|---|---|---|---|
| $\lambda_{s_i}$ | Average daily vaccine doses given at age group i | daily doses from data | **[38]** |
| $\phi_i$ | Susceptibility for age group i | 0.34 | 0.34 | 1 | 1 | 1.67 | 1.67 | **[39]** |
| $c_{ij}$ | Contacts per day | | **[32]** |
| | Reduction | 0.650043 — Phase I<br>0.759924 — Phase II<br>0.665048 — Phase III<br>0.77 — Phase IV | **Estimated** |
| $\beta$ | Probability of transmission | 2.30E-07 | **Estimated** |
| $\zeta$ | Increase in transmission from VOC | 1.5 | **Assumed** [17,18,19,20] |
| $\xi$ | Proportion of mild cases not adhering to self-isolation rule | 0.20002 | **Estimated** |
| $\alpha^{O,N}$ | average time in latent period | 1/4 $days^{-1}$ | **[40,41] (assumed for VOC)** |
| $\rho$ | Proportion of symptomatic individuals | 0.8 | **[42]** |
| $\gamma^{O,N}_{aR_i}$ | Recovery rate from asymptomatic infection | 0.16 $days^{-1}$ | **[43]** |
| $\gamma^{o}_{H_i}$ | Hospitalization rate of individuals in group I, infected with old variant | 0.0029 0.0005 0.0027 0.0108 0.0458 0.1006 — Phase I<br>0.0024 0.0020 0.0047 0.0113 0.0475 0.1194 — Phase II<br>0.0018 0.0015 0.0052 0.0156 0.0558 0.1371 — Phase III<br>0.0014 0.0005 0.0034 0.0101 0.0327 0.0773 — Phase IV | **Estimated** |
| $\gamma^{N}_{H_i}$ | | 0 0 0 0 0.2550 0.1133 — Phase I | |

| | | | | |
|---|---|---|---|---|
| | Hospitalization rate of individuals in group I, infected with VOC | 0.0027 0.0019 0.0066 0.0179 0.0911 0.1793 | Phase II | **Estimated** |
| | | 0.0010 0.0014 0.0070 0.0247 0.0708 0.1823 | Phase III | |
| | | 0.0019 0.0009 0.0057 0.0121 0.0426 0.1264 | Phase IV | |
| $\gamma_{mR_i}^{o}$ | Recovery rate of individuals in group I, mildly infected with old variant | 0.0991 0.0998 0.0992 0.0968 0.0865 0.0704 | Phase I | **Calculated** |
| | | 0.0993 0.0994 0.0986 0.0967 0.0860 0.0649 | Phase II | |
| | | 0.0995 0.0996 0.0985 0.0955 0.0840 0.0608 | Phase III | |
| | | 0.0996 0.0998 0.0990 0.0970 0.0902 0.0768 | Phase IV | |
| $\gamma_{mR_i}^{N}$ | Recovery rate of individuals in group I, mildly infected with VOC | 0.1000 0.1000 0.1000 0.1000 0.0250 0.0667 | Phase I | **Calculated** |
| | | 0.0992 0.0994 0.0981 0.0947 0.0732 0.0473 | Phase II | |
| | | 0.0997 0.0996 0.0980 0.0929 0.0798 0.0479 | Phase III | |
| | | 0.0994 0.0997 0.0983 0.0964 0.0872 0.0621 | Phase IV | |
| $\mu_{H_i}^{o}$ | Mortality rate from old variant | 0 0 0.0001 0.0013 0.0143 0.0834 | Phase I | **Estimated** |
| | | 0 0 0.0001 0.0012 0.0124 0.0522 | Phase II | |
| | | 0 0 0.0005 0.0008 0.0077 0.0407 | Phase III | |
| | | 0 0 0 0.0003 0.0007 0.0102 | Phase IV | |
| $\mu_{H_i}^{N}$ | Mortality rate from VOC | 0 0 0 0 0.1100 0 | Phase I | **Estimated** |
| | | 0 0 0.0005 0.0017 0.0238 0.1336 | Phase II | |
| | | 0 0 0.0002 0.0010 0.0092 0.0600 | Phase III | |
| | | 0 0 0 0 0.0015 0.0129 | Phase IV | |
| $\gamma_{HR_i}^{O}$ | Recovery rate of hospitalized individuals in group I, mildly infected with old variant | 0.1200 0.1200 0.1200 0.1196 0.1161 0.0972 | Phase I | **Estimated** |
| | | 0.1800 0.1800 0.1800 0.1794 0.1736 0.1532 | Phase II | |
| | | 0.1800 0.1800 0.1798 0.1796 0.1764 0.1607 | Phase III | |
| | | 0.1300 0.1300 0.1300 0.1299 0.1298 0.1272 | Phase IV | |
| $\gamma_{HR_i}^{N}$ | Recovery rate of hospitalized individuals in group I, mildly infected with VOC | 0.1200 0.1200 0.1200 0.1200 0.0900 0.1200 | Phase I | **Estimated** |
| | | 0.1800 0.1800 0.1798 0.1791 0.1678 0.1113 | Phase II | |
| | | 0.1800 0.1800 0.1799 0.1795 0.1756 0.1516 | Phase III | |
| | | 0.1300 0.1300 0.1300 0.1300 0.1296 0.1264 | Phase IV | |
| $\epsilon_{1_i}$ | Efficacy first dose for age group i | 0 \| 0.8 \| 0.8 \| 0.8 \| 0.7 \| 0.7 <br> reduced by 0.1 in lower efficacy scenario | | **Assumed [44]** |
| $\epsilon_{2_i}$ | Efficacy second dose for age group i | 0 \| 0.9 \| 0.9 \| 0.9 \| 0.8 \| 0.8 <br> reduced by 0.1 in lower efficacy scenario | | **Assumed [44]** |

| | | | |
|---|---|---|---|
| $\sigma$ | Average time to receive second dose | 1/112 days$^{-1}$ | [33] |
| $\omega_1$ | Average time to wane immunity after first dose | 1/120 days$^{-1}$ | **Assumed** |
| $\omega_2$ | Average time to wane immunity after second dose | 1/365 days$^{-1}$ | **Assumed** |
| $S_{0_i}$ | Susceptible individuals in age group I (initial value) | 283648 280541 901570 832208 546916 150982 | **Calculated** |
| $E_{0_i}$ | Exposed individuals in age group I (initial value) | Old: 158 301 1528 1169 553 264<br>VOC: 0 0 1 2 2 1 | **Calculated** |
| $A_{0_i}$ | Asymptomatic individuals in age group I (initial value) | Old: 46 78 340 300 152 61<br>VOC: 0 0 0 0 0 1 | **Calculated** |
| $I_{m0_i}$ | Symptomatic individuals in age group I (initial value) | Old: 340 529 2378 1936 877 318<br>VOC: 0 0 0 0 0 0 | **Calculated** |
| $H_{0_i}$ | Hospitalized individuals in age group I (initial value) | Old: 4 4 44 121 236 208<br>VOC: 0 0 0 0 0 0 | **Calculated** |
| $D_{0_i}$ | Deceased individuals in age group I (initial value) | Old: 1 0 4 79 527 1351<br>VOC: 0 0 0 0 0 0 | **Calculated** |
| $R_{0_i}$ | Recovered individuals in age group I (initial value) | Old 2735 4637 21635 17060 8010 3588<br>VOC 0 0 0 0 0 0 | **Calculated** |

| **SENSITIVITY ANALYSIS PARAMETERS** | | |
|---|---|---|
| **PARAMETER** | **DEFINITION** | **RANGE (uniform distribution)** |
| $\sigma$ | Rate at which second dose is distributed | [1/112, 1/21] |
| $\lambda_2$ | Daily doses age group 10-19 | [500, 2719] |
| $\lambda_3$ | Daily doses age group 20-39 | [1624, 8559] |
| $\lambda_4$ | Daily doses age group 40-59 | [2312, 8714] |
| $\lambda_5$ | Daily doses age group 60-79 | [599, 2702] |
| $\lambda_6$ | Daily doses age group 80+ | [319, 900] |

#### *Proportion of VOC cases*

To capture the increasing trend of cases from VOC, we defined a time-dependent function ($b(t)$) following a sigmoid function. Fig. A1 shows the proportion of cases from VOC from data (red circles) and the function used to reproduce their trend (blue curve). According to data up to May 19, 2021 the proportion of cases from VOC in Toronto reached a maximum of 0.8 by May 11, 2021. Hence, we consider 80% to be the maximum of cases generated by the new variant.

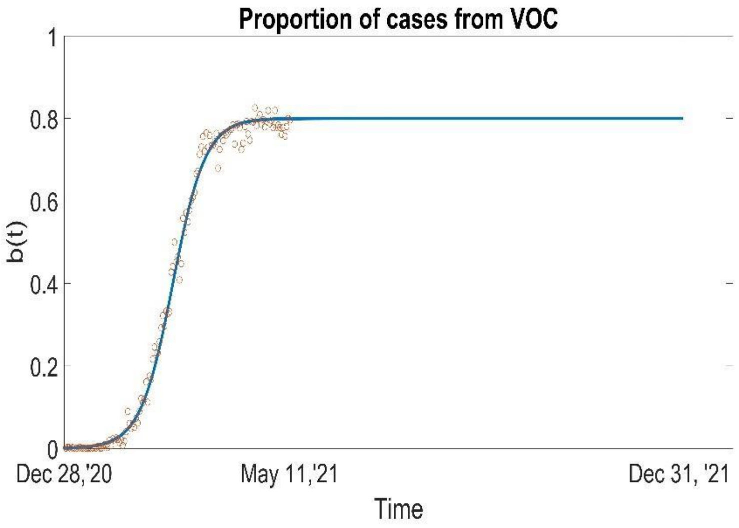


**Fig. SI1:** Sigmoidal function describing the growth of proportion of cases from VOC in Toronto. Scatter plot represents the proportion of VOC cases in Toronto from December 28, 2020 to May 11, 2021.

### *Data fitting*

To calibrate the model's parameters, we employed the Least Squared Method (LSM) using cumulative and daily cases and deaths, and hospitalizations (Figure SI2)

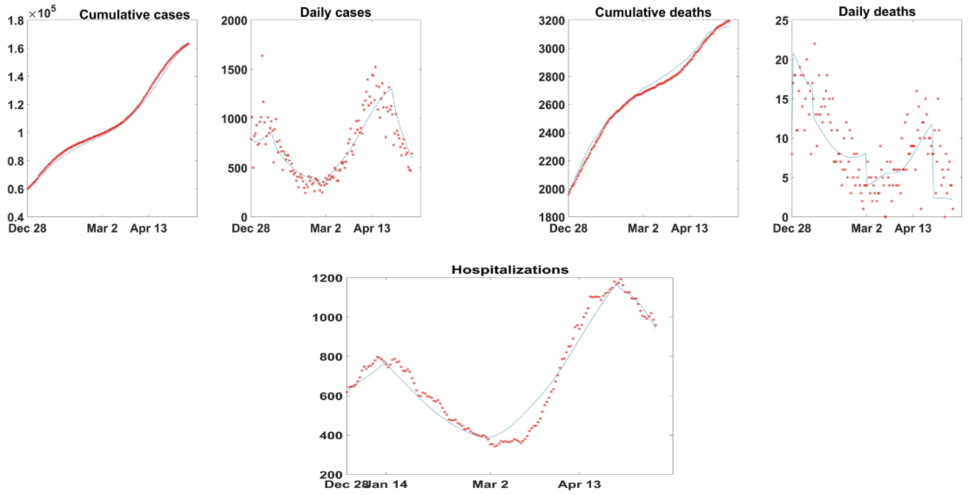


**Figure SI2:** Parameters' calibration using Least Square Method. We used cumulative and daily cases and deaths, and hospitalizations between December 28, 2020 and May 19, 2021.

### *Permutations of model's analysis*

All the scenarios used for the projections are shown in Figure SI3. Each scenario is described by taking one element in each column.

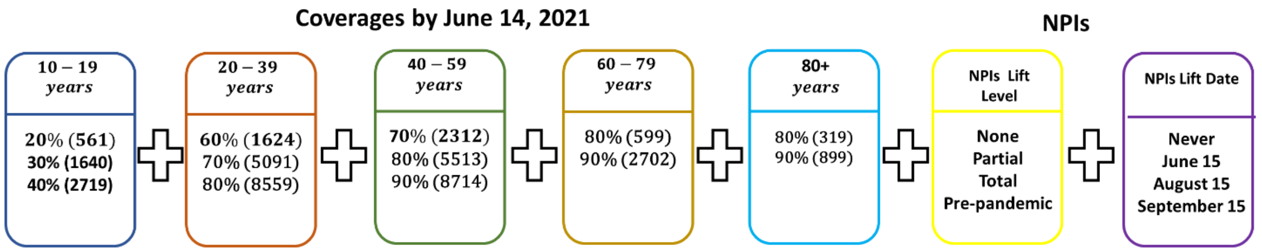


| Base line for analysis | | | | | | |
|---|---|---|---|---|---|---|
| **10-19 years** | **20-39 years** | **40-59 years** | **60-79 years** | **80 + years** | **NPIs lift level** | **NPIs lift date** |
| **20%** | **60%** | **70%** | **80%** | **80%** | **None** | **Never** |

**Figure SI3:** Outward-facing model coverages and base line for model's analysis. All these coverages are reached by June 14, 2021. In brackets, we report the daily doses. Each scenario is described by taking one element in each column.

### *Contact matrix*

We used the total contact matrix from a recent Canadian study [32]. However, the age groups used in this study were defined by a 5-year band from 0 to 80+. Our model is using larger age groups, then it was necessary to aggregate the original contact matrix in less groups.

Let's define $P_j$ the population size of age group $j \in \{1,2,3, \dots 17\}$, where $1 = 0 - 4\, years, 2 = 5 - 9\, years, \dots, 17 = 80 +$ years. To better approximate the contact rates, we calculated, from the original 17x17 matrix ($M_{ij}$), the total contacts that an age group has with all the other age groups. To obtain this, we multiplied all the age groups by their own population size, i.e. $m_{ij} \times P_j$ . Then, to aggregate some age groups, we averaged the total contacts as follows:

- For same ages belonging to new aggregation: we summed up the diagonal entries of the submatrix related to the age groups to aggregate and the average of the mixed contacts $\left(\hat{c}_{ii} = \sum m_{ii} + \sum \frac{m_{ij}+m_{ji}}{2}\right)$. For example, the new contact of the aggregated group 0-9, given by group 1 and 2, will be $m_{11} + m_{22} + \frac{m_{12}+m_{21}}{2}$
- For different ages aggregation: we summed up the average of the mixed contacts $\left(\hat{c}_{ij} = \sum \frac{m_{ij}+m_{ji}}{2}\right)$. For example, the new contact of the aggregated group 0-9 and 10-19, given by group 1, 2, 3 and 4, will be $\frac{m_{13}+m_{31}}{2} + \frac{m_{14}+m_{41}}{2} + \frac{m_{23}+m_{32}}{2} + \frac{m_{24}+m_{42}}{2}$

Once we reduced the total contacts into a smaller matrix, we re-parametrized each entry of the new age group dividing the obtained contacts by the population size of the aggregate age group (i.e., $c_{ij} = \hat{c}_{ij} / \sum P_j$ ). Table A2 represents the compacted matrix.

**Table SI3:** Contact matrix

| | | Age participants | | | | | |
|---|---|---|---|---|---|---|---|
| | | **0-9** | **10-19** | **20-39** | **40-59** | **60-79** | **80+** |
| **Age contacts** | **0-9** | 2.59 | 0.54 | 0.51 | 0.67 | 0.21 | 0.04 |
| | **10-19** | 0.56 | 3.13 | 0.66 | 0.86 | 0.21 | 0.10 |
| | **20-39** | 1.88 | 2.35 | 2.52 | 3.06 | 1.29 | 0.65 |
| | **40-59** | 1.88 | 2.33 | 2.33 | 2.10 | 1.50 | 1.10 |
| | **60-79** | 0.40 | 0.39 | 0.66 | 1.00 | 1.25 | 1.04 |
| | **80+** | 0.02 | 0.05 | 0.09 | 0.20 | 0.28 | 0.58 |

### *RESULTS*

### *Reproduction number $R_c$*

**Figure SI4**: Contour plots of $R_c$ assuming that the following coverages reached for age groups 10-19, 60-79 and 80+ years are 20%, 80%, and 90%, respectively, when the NPIs level reopening is (A) partial, (B) total an (C) pre-pandemic. As expected, as the vaccination coverage increases, the values of the reproduction number decrease. Also, we observe that with the lowest reopening level, to reduce the reproduction number below 1, it is sufficient to vaccinate age groups 20-39 and 40-59 years above 60% and 62%, respectively. On the other hand, a relaxation of NPIs and increase in contacts as in NPIs partial reopening, the $R_c$ will always be greater than 1. Similar results, but higher $\boldsymbol{R_c}$, are shown with NPIs pre-pandemic reopening (C).

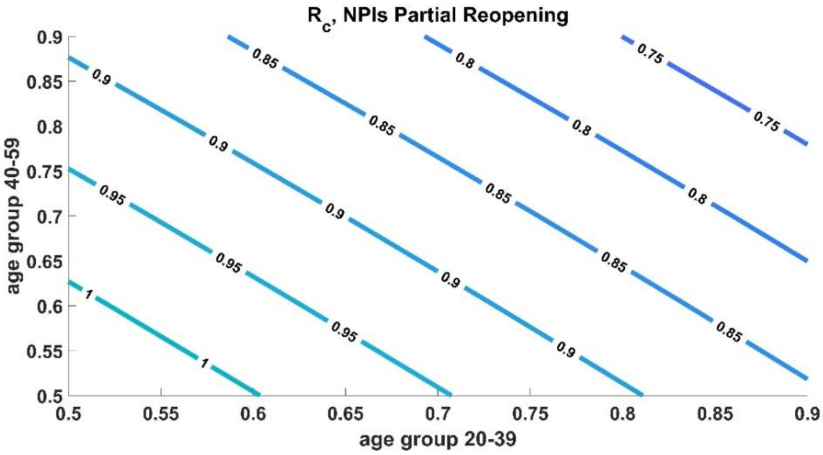


A

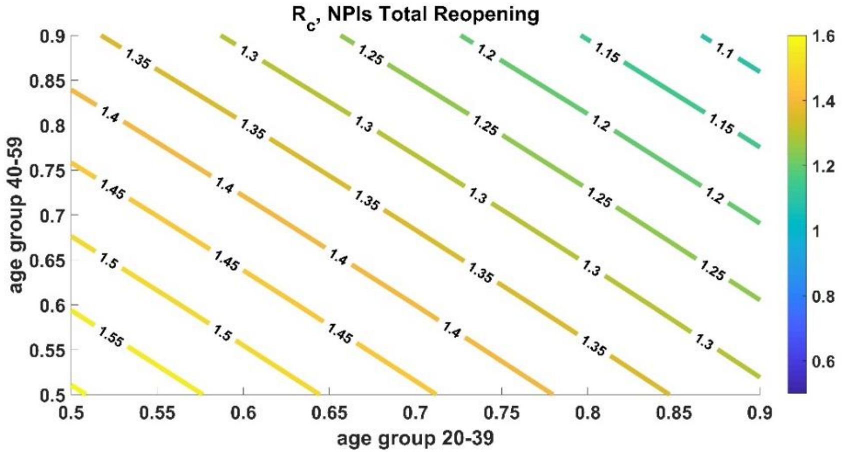


B

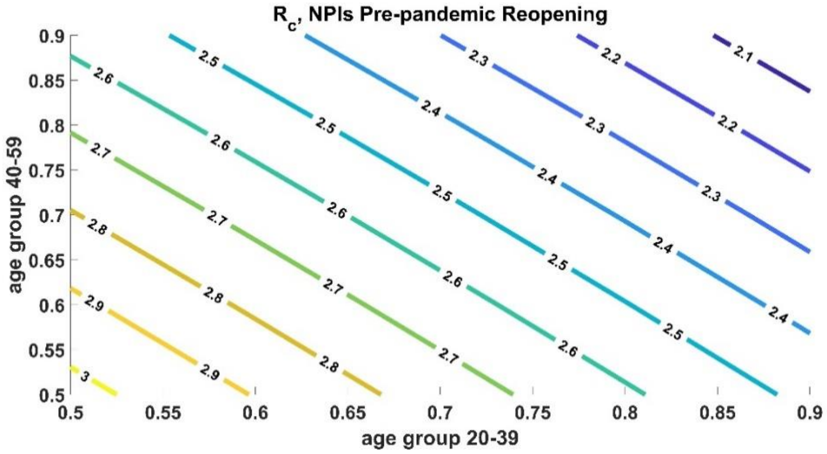


C

### *Projections*

**Table SI4** : Percentage change of cumulative deaths with respect to the baseline with respect to the base line NPIs no reopening in SI Figure SI3 with partial, total and pre-pandemic reopening in August and September, when age groups 10-19, 60-79 and 80+ reached coverages 20%, 80%, 90%. The second dose is given at a rate of 1/112 days$^{-1}$.

<table>
<tr><th colspan="8">Projected percentage change of <u>cumulative cases</u> with respect to the base line NPIs no reopening in SI Figure SI3</th></tr>
<tr><td colspan="2" rowspan="3"></td><td colspan="6">If reopen in AUGUST</td></tr>
<tr><td colspan="6">20-39 years<br>coverage by June 14, 2021</td></tr>
<tr><td colspan="3">60%</td><td colspan="3">80%</td></tr>
<tr><td colspan="2">NPI's Level of reopening</td><td>Partial</td><td>Total</td><td>Pre-pandemic</td><td>Partial</td><td>Total</td><td>Pre-pandemic</td></tr>
<tr><td rowspan="3">40-59 years coverage by June 14, 2021</td><td>70%</td><td>4.15</td><td>202</td><td>1178</td><td>1.51</td><td>71.34</td><td>1175</td></tr>
<tr><td>80%</td><td>2.75</td><td>138.4</td><td>1175</td><td>0.93</td><td>45</td><td>1169</td></tr>
<tr><td>90%</td><td>1.77</td><td>88</td><td>1169</td><td>0.49</td><td>27.6</td><td>1159</td></tr>
<tr><td colspan="2" rowspan="3"></td><td colspan="6">In reopen in SEPTEMBER</td></tr>
<tr><td colspan="6">20-39 years<br>coverage by June 14, 2021</td></tr>
<tr><td colspan="3">60%</td><td colspan="3">80%</td></tr>
<tr><td colspan="2">NPI's Level of reopening</td><td>Partial</td><td>Total</td><td>Pre-pandemic</td><td>Partial</td><td>Total</td><td>Pre-pandemic</td></tr>
<tr><td rowspan="3">40-59 years coverage by June 14, 2021</td><td>70%</td><td>-1.5</td><td>33.4</td><td>1167</td><td>-1.58</td><td>10.67</td><td>1058</td></tr>
<tr><td>80%</td><td>-1.55</td><td>20.8</td><td>1130</td><td>-1.61</td><td>6.3</td><td>977</td></tr>
<tr><td>90%</td><td>-1.59</td><td>12.36</td><td>1075</td><td>-1.64</td><td>3.7</td><td>871</td></tr>
</table>

**Figure SI5:** Hospitalizations with partial reopening in August (A) if 40-59 is vaccinated 70%-00%, 20-39 60%, 80% and 10-19, 60-79 and 80+ reached coverages 20%, 80%, 90%. The second dose is given at a rate of 1/112 days$^{-1}$.

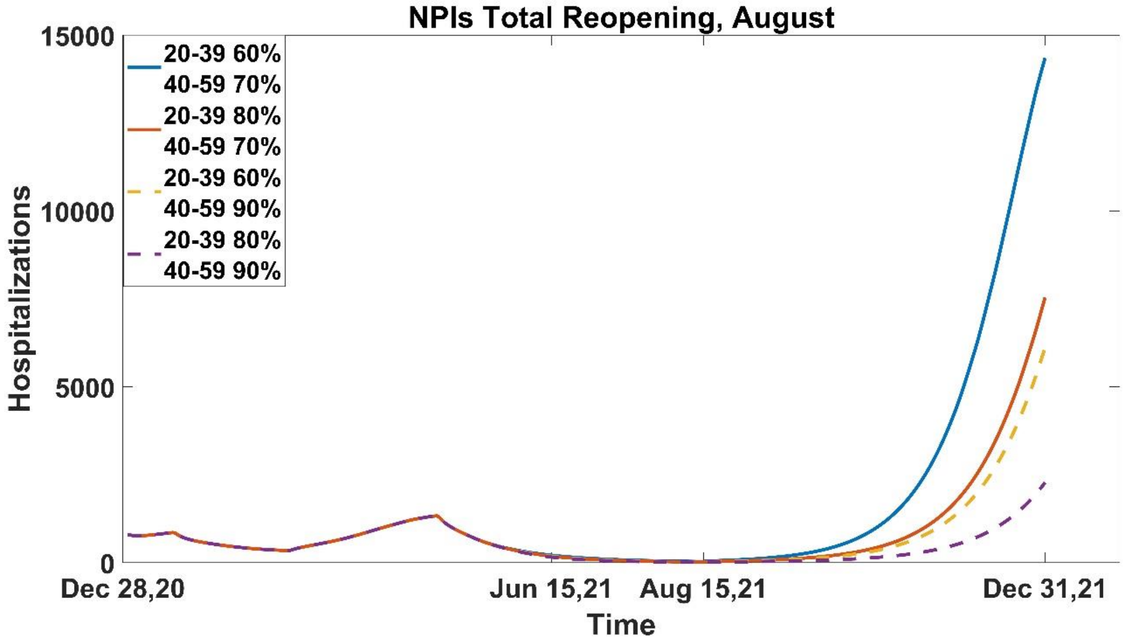


**Table SI5:** Percentage change of cumulative cases and deaths with respect to the baseline with respect to the base line NPIs no reopening in SI Figure SI3, reducing efficacy by 10%, with partial, total and pre-pandemic reopening when age groups 10-19, 60-79 and 80+ reached coverages 20%, 80%, 90%.

| **Projected percentage change of cumulative deaths with respect to the base line NPIs no reopening in SI Figure SI3 with reopening in September and efficacy reduced by 10%** | | | | | | | |
|---|---|---|---|---|---|---|---|
| | | **20-39 years coverage by June 14, 2021** | | | | | |
| | | **60%** | | | **80%** | | |
| **NPI's Level of reopening** | | **Partial** | **Total** | **Pre-pandemic** | **Partial** | **Total** | **Pre-pandemic** |
| **40-59 years coverage by June 14, 2021** | **70%** | **-1.51** | 48.56 | 1220 | **-1.58** | 18.58 | 1152 |
| | **80%** | **-1.55** | 32.12 | 1192 | **-1.61** | 12.27 | 1100 |
| | **90%** | **-1.59** | 21.6 | 1160 | **-1.63** | 7.54 | 1017 |

**Table SI6**: Percentage change of cumulative cases and deaths with respect to the base line NPIs no reopening in SI Figure SI3 with partial, total and pre-pandemic reopening in September and second dose given after 21 or 50 days. Age groups 10-19, 60-79 and 80+ are assumed to reach coverages 20%, 80%, 90% by mid June. Par.= partial; Tot.= total; Pre-pan.= pre-pandemic..

| **Projected percentage change of cumulative deaths with respect to the base line NPIs no reopening in SI Figure SI3 with reopening in September** | | | | | | | | | | | | | |
|---|---|---|---|---|---|---|---|---|---|---|---|---|---|
| | | **21 days Between dose 1 and dose 2** | | | | | | **50 days Between dose 1 and dose 2** | | | | | |
| | | **20-39 years coverage by June 14, 2021** | | | | | | **20-39 years coverage by June 14, 2021** | | | | | |
| | | **60%** | | | **80%** | | | **60%** | | | **80%** | | |
| **NPI's Level of reopening** | | **Par.** | **Tot.** | **Pre-pan.** | **Par.** | **Tot.** | **Pre-pan.** | **Par.** | **Tot.** | **Pre-pan.** | **Par.** | **Tot.** | **Pre-pan.** |
| **40-59 years coverage by June 14, 2021** | **70%** | **--1.76** | 3.8 | 719 | **-1.81** | **0.30** | 370 | **-1.61** | 11.8 | 942 | **-1.67** | 2.8 | 722 |
| | **80%** | **-1.8** | 1.64 | 577 | **-1.83** | **-0.21** | 216 | **-1.65** | 6.76 | 868 | **-1.7** | **1.32** | 567 |
| | **90%** | **-1.82** | **0.47** | 414 | **-1.85** | **-0.48** | 110 | **-1.68** | 3.54 | 759 | **-1.72** | **0.52** | 400 |

**Figure SI6:** Hospitalizations with total reopening in September if 40-59 is vaccinated 70%-00%, 20-39 60%, 80% and 10-19, 60-79 and 80+ reached coverages 20%, 80%, 90% and if the second dose is given at a rate of (A) 1/21 days$^{-1}$ or (B) 1/50 days$^{-1}$.

**A**

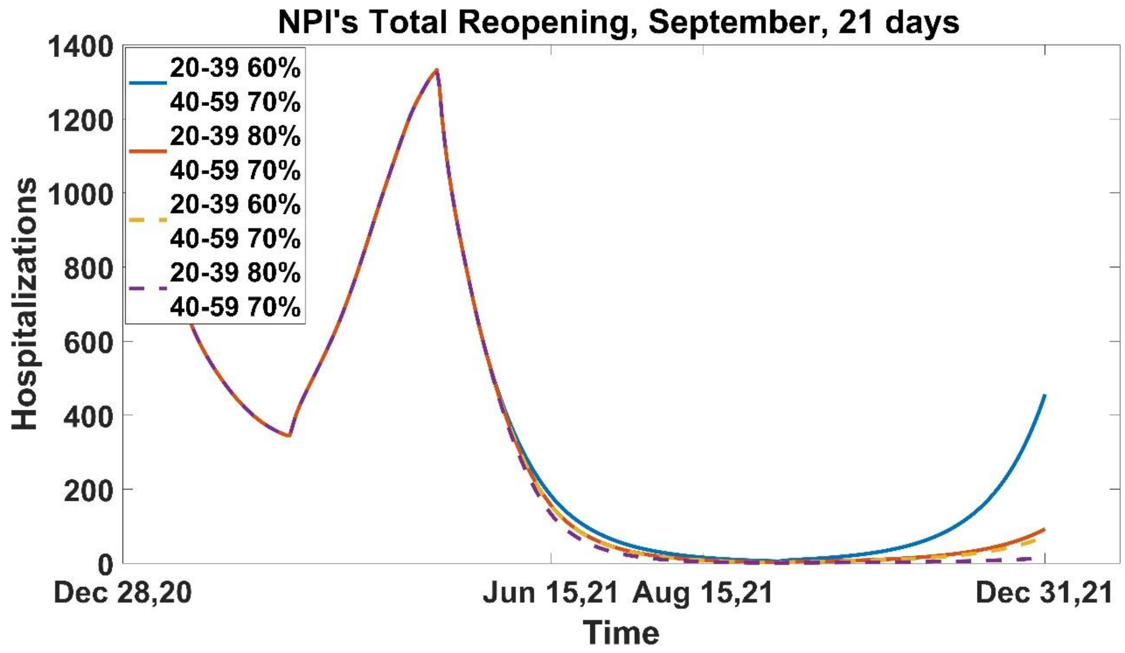


**B**

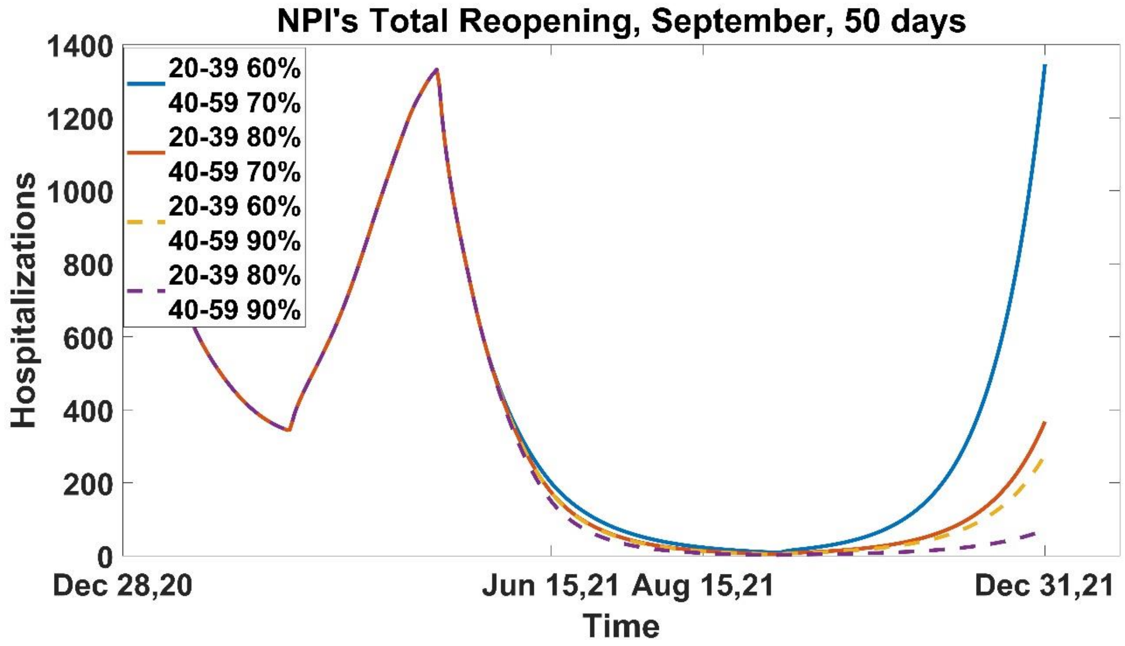


### *Sensitivity Analysis*

Using the Latin Hypercube Sampling/Partial Rank Correlation Coefficient (*LHS*/*PRCC*) we conducted sensitivity analysis on the parameters related to vaccination, such as level of coverage, waning immunity, minimum time to reach a certain level of vaccination, as well as infection-related parameters.

**Table SI7:** PRCC on cumulative cases and deaths.

| **SENSITIVITY ANALYSIS** | | | | |
|---|---|---|---|---|
| **PARAMETERS** | **DEFINITION** | **PRCC** | | |
| | | CASES | DEATHS | HOSPITALIZATION (50 days after reopening in June) |
| $\sigma$ | Rate at which second dose is distributed | **-0.9503** | **-0.9503** | **-0.9739** |
| $\lambda_2$ | Daily doses age group 10-19 | -0.08731 | -0.08731 | -0.05824 |
| $\lambda_3$ | Daily doses age group 20-39 | **-0.8041** | **-0.8041** | **-0.862** |
| $\lambda_4$ | Daily doses age group 40-59 | **-0.7653** | **-0.7653** | **-0.8558** |
| $\lambda_5$ | Daily doses age group 60-79 | -0.163 | -0.163 | -0.3133 |
| $\lambda_6$ | Daily doses age group 80+ | 0.02592 | 0.02592 | -0.01986 |

Table SI7 shows the PRCCs of the sampled parameters $\lambda_i$, $i \in \{2,3,4,5,6\}$, and $\sigma$ , the daily doses in age group i, and the rate of receiving the second dose, respectively, on the cumulative cases and deaths. We observe that the age groups 3 and 4, namely, 20-39 and 40-59 years present the highest PRCC among the daily doses, suggesting that an increased vaccine coverage of these age groups leads to the largest reduction in cases and deaths. Moreover, $\sigma$ is negatively correlated to cases and deaths, suggesting that if this rate is small, hence the time between doses is longer, cases and deaths will increase. Similar results are visible for the hospitalizations reported 50 days after reopening in June.
